# Knowledge, attitude, and perceptions towards the 2019 Coronavirus Pandemic: A bi-national survey in Africa

**DOI:** 10.1101/2020.05.27.20113951

**Authors:** Elnadi Hager, Ismail A. Odetokun, Obasanjo Bolarinwa, Ahmed Zainab, Ochulor Okechukwu, Al-Mustapha A. Ibrahim

**Affiliations:** Infectiologie et Santé Publique, Institut National de la Recherche Agronomique, Nouzilly, France; Department of Veterinary Public Health and Preventive Medicine, Faculty of Veterinary Medicine, University of Ilorin, Kwara State, Nigeria; School of Nursing & Public Health Medicine, College of Health Sciences, University of KwaZulu - Natal, Durban, South Africa; Department of Veterinary Microbiology, Faculty of Veterinary Medicine, Cairo University, Egypt; Department of Global Health, Unit of Functional Genetics of Infectious Diseases, Institute Pasteur, France; Department of Food Hygiene and Environmental Health, Faculty of Veterinary Medicine, University of Helsinki, Finland; Department of Veterinary Services, Kwara State Ministry of Agriculture and Rural Development, Ilorin, Nigeria

**Keywords:** Knowledge, attitude, perception, COVID-19, Nigeria, Egypt

## Abstract

The current Coronavirus (COVID-19) pandemic has changed and impacted lives on a global scale since its emergence and spread from China in late 2019. It has caused millions of infections, and thousands of deaths worldwide. However, the control of this pandemic still remains unachievable in many African countries including Egypt and Nigeria, despite the application of some strict preventive and control measures. Therefore, this study assessed the knowledge, attitude and perceptions of Egyptians and Nigerians towards COVID-19 pandemic.

A total of 1437 respondents were included in this preliminary cross-sectional survey. The mean knowledge score was 14.7±2.3. The majority of the respondents (61.6%) had a satisfactory knowledge of the disease. Age (18-39 years), education (College/bachelors) and background of respondents were factors influencing knowledge levels. The attitude of most respondents (68.9%) towards the preventive measures was satisfactory with an average attitude score of 6.9 ± 1.2. The majority of the respondents (96%) practiced self-isolation and social-distancing but only 36% follow all health recommendations. The perception of most respondents (62.1%) on the global efforts at controlling the virus and preventing further spread was satisfactory with an average score of 10.9 ± 2.7. A satisfactory knowledge of COVID-19 was significantly associated with good attitude and perceptions (p < 0.001) of respondents. Only 22% of the respondents were satisfied with their country’s handling of the pandemic.

It is imperative that to avoid Africa being the next epicenter of the pandemic. Governments need to strengthen health systems, improve their surveillance activities in detecting cases, and effectively apply standard infection prevention and control measures.

## Introduction

The World Health Organization (WHO), on December 31, 2019, received a report of the presence of unknown causes of pneumonia disease in Wuhan, China (1). Later, this disease was defined as a novel Coronavirus disease and further declared as a public health emergency of international concern by January 30, 2020 (2). The novel virus was renamed by the International Committee on Taxonomy of Viruses, as severe acute respiratory syndrome coronavirus 2 (SARS-CoV-2) that causes the 2019 Coronavirus disease (COVID-19) (3; 4). COVID-19 is caused by a single stranded RNA virus belonging to the Coronaviridae family (5). This disease is similar to the previously emerged SARS-CoV and the Middle East respiratory syndrome Coronavirus (MERS-CoV) (6). Still, unlike these, its outbreaks have taken a global pandemic course. Since the first report of the confirmed cases of the COVID-19 in Wuhan, China (1;7), the world has witnessed severe unprecedented mortality and morbidity due to this disease resulting in serious public health emergencies. Infection by SARS-CoV-2 in humans occurs mainly through air droplets, close contact with infected persons, especially mucus membranes secretions from nose, mouth, or eyes, contaminated surfaces and some studies suggest digestive tract transmission (8; 9).

Despite the level of advancement in health systems in developed countries like the United States, the United Kingdom, France, Italy, and Spain, they appeared to be the worst hit with the epidemic curves still rising (10). No proven treatments or vaccines are available to control COVID-19 and thus pose a significant threat to health care delivery. To flatten the curves, most nations, including African countries, have applied strict prevention and control measurements to curb the disease including regulations such as general lockdown, obligatory home quarantine, ban on public gatherings, international flights restrictions and raising awareness on proper hand wash, hygiene and sanitation as well as social distancing (11).

The rate of infection due to COVID-19 on the African continent is on the increase, especially in Egypt in the north and Nigeria in the west. As of May 14, there were 72,336 confirmed cases, 2475 deaths, and 25,270 recoveries due to COVID-19 in Africa (12), with approximately 22% of these cases from Egypt and Nigeria alone. To stop this pandemic, it is imperative to institute effective infection prevention and control practices globally, nationally, and at the community level. Consequently, it is urgent to understand the public knowledge, reactions, adherence to, and acceptance of such measures that affect their daily life in several ways, especially psychologically, socially, and physically. This could be achieved through knowledge, attitude, and practice (KAP) studies. The information generated from such studies, in addition to comprehensive reviews and recommendations, could help in the fight against COVID-19 and similar future threats (13; 14).

In this study, we investigated the public response from two African countries (Egypt and Nigeria), towards the COVID-19 outbreak. This is a first report on the knowledge, attitude, and perceptions of participants with a scope covering more than one African country. Findings from this study would contribute to the global efforts to control the COVID-19 pandemic.

## Materials and Methods

### Study design

This study was conducted in April 2020 using an online cross - sectional survey of respondents from two African countries – Egypt and Nigeria. Egypt and Nigeria currently rank high in the number of confirmed cases for COVID-19 from the northern and western regions of Africa respectively.

### Study participants, sample size and sampling

The targeted respondents from both countries include adults >17 years of all educational levels, including both medical and non-medical backgrounds. To calculate the sample size for this survey, we hypothesized that the percentage frequency of outcome factor in the population *(p*) is 50% with a design effect of 1 at a confidence level of 99.9%. A sample size of 1083 was calculated using the Open Source Epidemiologic Statistics for Public Health (OpenEpi), v.3.01 (updated 2013/04/06). To make up for non-response, 30% non-contingency was added. Thus, a minimum of 1,408 were targeted to be obtained from both countries. Since, Nigeria has a population more than twice of Egypt, the respondents were sampled in at least a ratio of 1 (Egypt): 3 (Nigeria). A preliminary analysis of 1,437 respondents (Nigeria - 1,132; Egypt - 305), recruited using a convenient sampling method, was conducted to assess their knowledge, attitude, and perceptions towards the pandemic. The online survey was carried out between April and May 2020. Due to the spread of the COVID-19 pandemic and the lockdown policy enforced in both countries, respondents were reached via emails and social media platforms such as WhatsApp and Facebook messenger simultenously in both countries. Initially, respondents from major cities, Lagos/Ilorin (Nigeria) and Cairo/Alexandria (Egypt) were recruited before the questionnaire administration spread to participants from other major cities and towns across the two countries. The online web-based survey was anonymous and administered in the official languages (Arabic and English) of both countries.

### Ethical considerations

The Kwara State Ministry of Education, Ilorin, Nigeria (reference number: DE/PRIM/96/VOL.1/130) granted approval for the conduct of this study. This approval suffices for the surveys in both countries. Participation was anonymous and voluntary. Informed consent was sought from the respondents and participants could withdraw from the survey at any time in line with stipulations of the World Medical Association Declaration of Helsinki Ethical principles (15).

### Questionnaire design

We designed a structured questionnaire using google forms (Alphabet Inc., California, USA). The survey tool is available online (https://forms.gle/h649kakEzLAXcpYo7). The questionnaire was pre-validated by three independent reviewers, and a pre-test study was conducted with 20 respondents from Nigeria. The responses from the pre-test were not included in the analyzed data but used to improve upon the quality of the questionnaire. The questionnaire consisted of 5 parts: a). Demography of respondents, b). Knowledge of Coronavirus (COVID-19), c). Attitude towards preventive measures, d). Perception of global response, and e). Community response to the pandemic. The survey was designed as a quiz. We provided the correct answers to all questions wrongly answered by the respondents as a feedback. All questions and responses were based on the latest recommendations by the WHO (1; 3). Section B tested their knowledge of/focused on disease spread, symptoms, incubation period, and how to limit infection. Section C evaluated their attitude towards preventive measures by focusing on questions related to hand hygiene, wearing face masks, and social distancing. Sections D and E assessed their perception of global and community response efforts to the pandemic with particular emphasis on ways to prevent future occurrence of such outbreaks.

### Data analysis

Data were summarized using Microsoft Excel 2019 and analyzed utilizing the Statistical Package for the Social Sciences (SPSS) software, v.22, and the OpenEpi. To summarize the obtained data, the demographic characteristics of respondents were subjected to descriptive statistics (frequency and proportions). To assess knowledge, attitude, and perception levels of respondents, a numeric scoring pattern was used, and outcome (dependent) variables – knowledge, attitude, and perception – were computed (16). These outcome variables were further categorized as binary (satisfactory or unsatisfactory) based on cut-off (mean scores) marks (Table 1). Respondents receiving scores greater than the mean scores for knowledge (14.7±2.3), attitude (6.9 ± 1.2), and perception (10.9 ± 2.7) were deemed to be satisfactory responses and vice versa. Chi-square test was used to test for association between independent variables (demographics) and outcome variables (knowledge, attitude and perception) at a 95% confidence interval with significant variables (p < 0.05) subjected to a logistic regression model.

## Results

### Respondent demographics

A total of 1437 respondents were included in this preliminary survey. Most respondents (83.3%, n = 1197/1437) were between the ages of 18 - 39 years. Similarly, the majority of the respondents (84.9%, n = 1220/1437) has a bachelor/master’s degree (Table 2). Respondents with a scientific/medical background accounted for 59.3% of the responses (n = 852/1437).

**Table 1:** Description of scores obtained by respondents (n = 1437)

| Outcome variables | Maximum obtainable scores | Scores received by respondents |  | Mean ± SD | Satisfactory n (%) | Unsatisfactory n (%) |
| --- | --- | --- | --- | --- | --- | --- |
|  |  | Minimum score | Maximum score |  |  |  |
| Knowledge | 20 | 5 | 18 | 14.7±2.3 | 885 (61.6) | 552 (38.4) |
| Attitude | 9 | 2 | 9 | 6.9 ± 1.2 | 990 (68.9) | 447 (31.1) |
| Perception | 17 | 1 | 16 | 10.9 ± 2.7 | 892 (62.1) | 545 (37.9) |

Cut-off marks = mean scores; Satisfactory scores = scores > mean score obtained by respondents; SD - standard deviation.

**Table 2:** Demographics of respondents from Nigeria and Egypt used in this preliminary study (n = 1437).

| Variable | Number of respondents (%) |
| --- | --- |
| Age (years) |  |
| 18 – 29 | 706 (49.1) |
| 30 – 39 | 491 (34.2) |
| 40 – 49 | 168 (11.7) |
| 50 – 59 | 51 (3.5) |
| 60 – 69 | 20 (1.4) |
| >69 | 1 (0.1) |
| Gender |  |
| Male | 754 (52.5) |
| Female | 677 (47.1) |
| Prefer not to say | 6 (0.4) |
| Education |  |
| No formal education | 2 (0.1) |
| High School | 60 (4.2) |
| College (Bachelor) | 897 (62.4) |
| Masters | 323 (22.5) |
| Ph.D. | 91 (6.3) |
| Others | 64 (4.5) |
| Background |  |
| Non-Scientific/Non-Medical | 585 (40.7) |
| Scientific/Medical | 852 (59.3) |
| Nationality |  |
| Nigeria | 1132 (78.8) |
| Egypt | 305 (21.2) |

% - percentage.

### Knowledge, attitude and perception of respondents towards COVID-19

#### Knowledge

The mean knowledge score was 14.7±2.3, from a maximum obtainable score of 20 (Table 1). Most respondents (61.6%, n = 885/1437) had satisfactory knowledge of the disease, and the internet was the main source of information for most respondents (83.7%, n = 1204/1437). Moreover, most (78%, n = 1127/1437) of the respondents knew that COVID-19 was different from common cold. The majority of the respondents knew that it is possible to have asymptomatic COVID-19 positive patients. Most respondents also knew that most symptoms appear between 1-14 days. Most respondents also correctly identified several symptoms of COVID-19, knew how to kill (inactivate) the virus, and recognized the importance of handwash in reducing the chances of contracting the disease (Table s1). All of the independent variables (age, gender, level of education, background, and nationality) were significantly (p < 0.05) associated with the knowledge of respondents about COVID-19.

#### Attitude

The participants attitude towards COVID-19 was satisfactory as the mean attitude score was 6.9 ± 1.2, with a range of 2 to 9 (Table1). Most of the respondents (68.9%, n = 990/1437) had a positive attitude towards protective measures being advised by the WHO or their local health authorities (Table 1). Most respondents (>80%) valued the importance of proper hygiene, self-isolation, the use of face mask when going out, and the ideal distance between two people in curbing the spread of the virus (Table s2). Some of the respondents were bored, fearful, and anxious to return to the “new normal.” Due to the compulsory lockdown, which has psycho - socially affected the lifestyle of most Nigerians and Egyptians, people have adapted by following the social media platforms (84%, n = 1207/1437), among other means of changing.

#### Perception

Respondents (62.1%, n = 892/1437) had a positive perception of global efforts to control the pandemic (Table 1). Although most of the respondents (81%, n = 1163/1437) agreed with the compulsory lockdown to prevent the further spread of the disease, only 38.6% (n = 554/1437) believe that the government had done enough to protect its citizens. Most respondents (77%, n = 1110/1437) rated their country’s national COVID-19 response plan below average (1-3 on a scale of 5) (Table s3).

The satisfactory knowledge of the respondents had a positive impact (p <0.001) on their attitudes towards preventive measures and their perception of a community response to curb the spread of the virus (Table 3). Most respondents (>81%, n = 1163/1437) agreed that improved personal hygiene, reducing social contacts, and following their countries health recommendations are necessary to reduce disease burden and reduce person to person transmission. The majority of the respondents (66%, n = 945/1437) believed that we can prevent a future pandemic by reducing international travels (33%, n = 472/1437); establishing improved early alerts and global warning systems for infectious diseases (82%, n = 1175/1437) and improving disease surveillance in both human and animal health sectors (73%, n = 1044/1437) (Table s4).

**Table 3:**
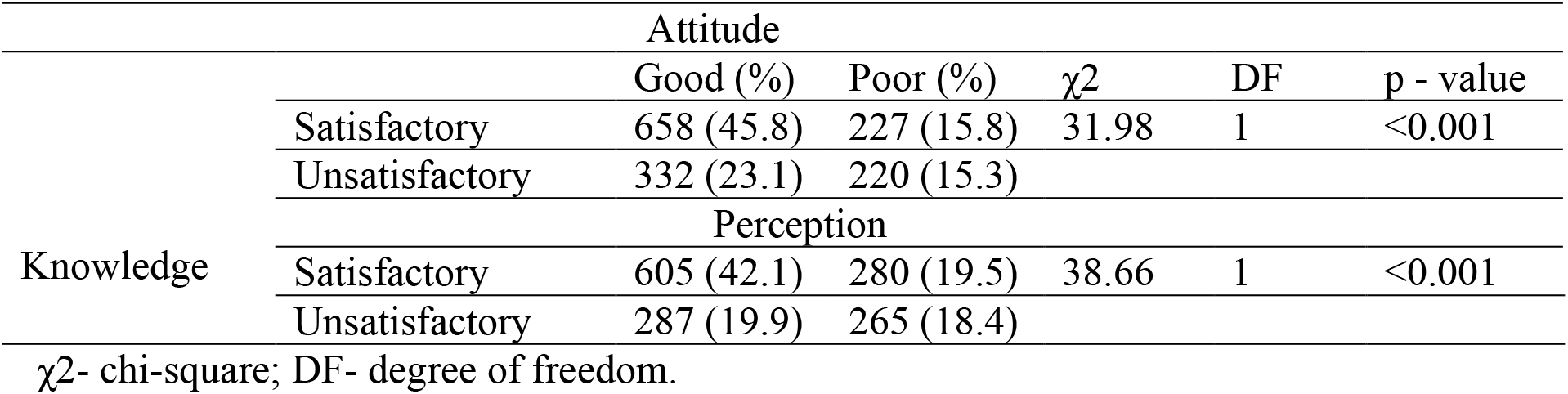
Test of association (Fischer’s exact test) between knowledge, attitude, and perception of respondents from Nigeria and Egypt (n = 1437).

### Demographic factors influence Knowledge, attitude and perception of respondents on COVID-19

Respondents within the 18 - 29 years age range were 1.4× (95%CI: 0.55 - 0.89; p = 0.004) more likely to be knowledgeable than other age groups. Respondents with a high school education were at least 4.7× (95% CI: 0.15 - 144.7; p = 0.73) more likely to have satisfactory knowledge about COVID-19 than those with no formal education. As expected, respondents with scientific or medical backgrounds were 1.4× (95% CI: 0.56 - 0.86; p < 0.001) more likely to be knowledgeable than those with non-scientific/non-medical background. Egyptians were 1.8× (95%CI: 0.43 – 0.74; p < 0.001) more likely to have more satisfactory knowledge than Nigerians (Table 4).

**Table 4.**
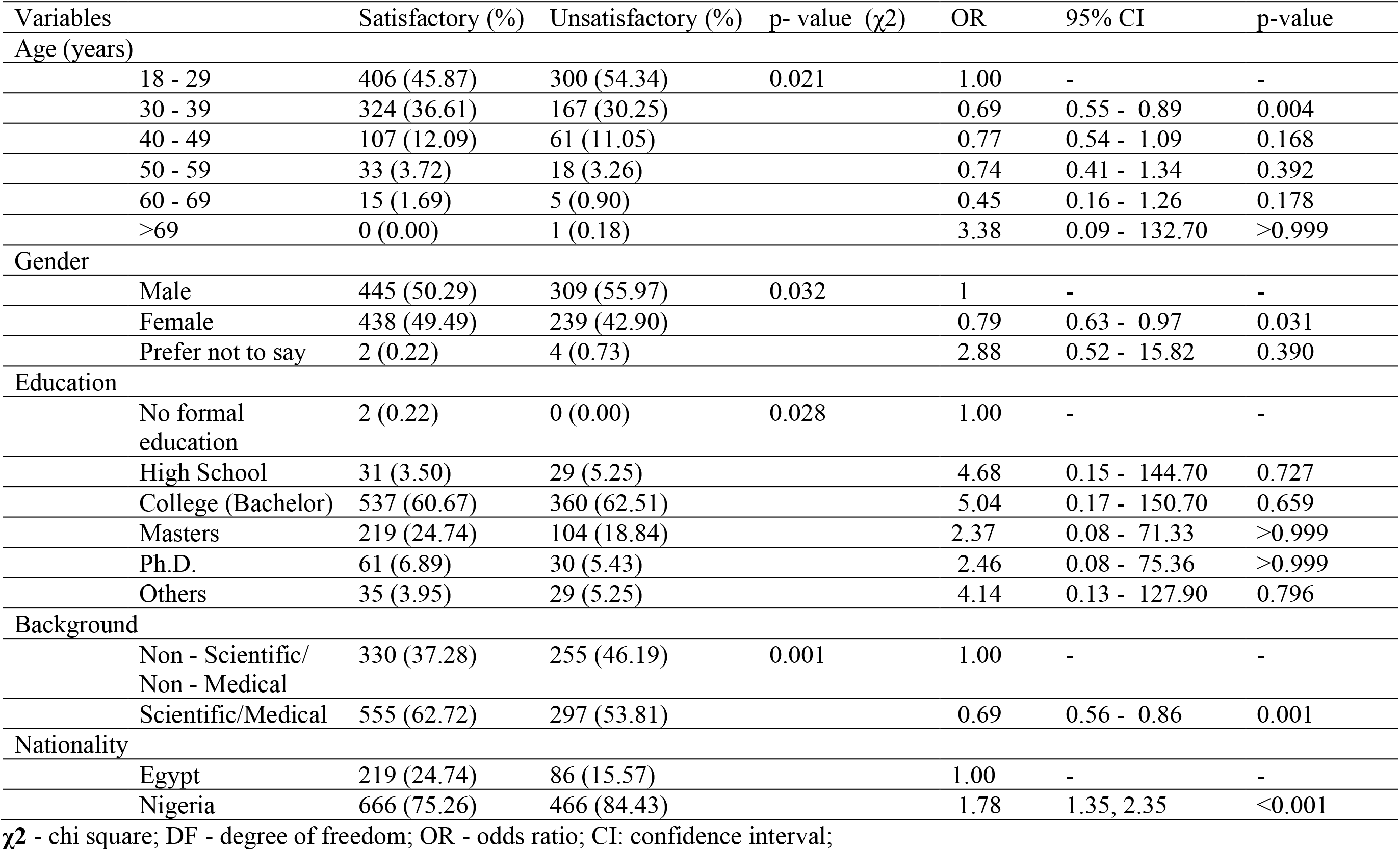
Analysis of demographic characteristics as factors influencing the knowledge levels of poultry farmers on antimicrobial in Kwara state.

The age, gender, level of education, background, and nationality had a significant impact on the attitude towards COVID-19. The older the respondents, the better their attitude towards the disease with an odds ratio ranging from 1.34 (95% CI: 1.06 - 1.74; p = 0.019) to 6.65 (95% CI: 0.17 - 206.9; p = 0.692). Female respondents were 1.59× (95% CI: 1.27 - 1.99; p < 0.001), more likely to have a positive attitude towards COVID-19 than males. Respondents of scientific/medical background were 1.6× (95% CI: 0.49 - 0.78; p <0.001) more likely to have better attitude than those with non-scientific/non-medical background. Nigerians were 11× (95% CI: 7.57 - 13.47; p <0.001) more likely to have a positive attitude than Egyptians (Table 5).

**Table 5.**
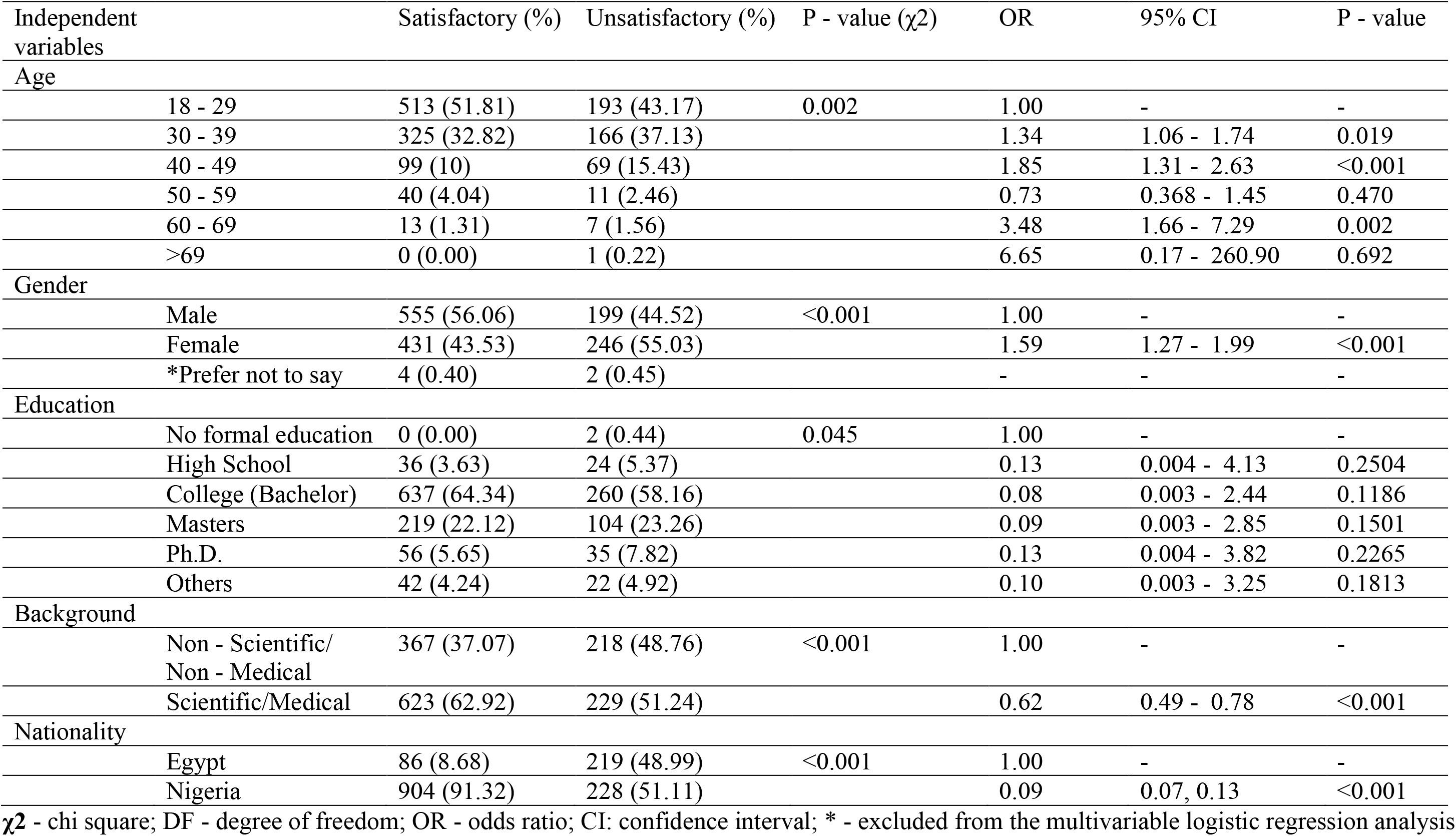
Analysis of demographic characteristics as factors influencing the attitude of respondents from Nigeria and Egypt towards COVID-19 pandemic.

| Independent variables | | Satisfactory (%) | Unsatisfactory (%) | P - value ($\chi^2$) | OR | 95% CI | P - value |
| --- | --- | --- | --- | --- | --- | --- | --- |
| Age |  |  |  |  |  |  |  |
|  | 18 - 29 | 513 (51.81) | 193 (43.17) | 0.002 | 1.00 | - | - |
|  | 30 - 39 | 325 (32.82) | 166 (37.13) |  | 1.34 | 1.06 - 1.74 | 0.019 |
|  | 40 - 49 | 99 (10) | 69 (15.43) |  | 1.85 | 1.31 - 2.63 | <0.001 |
|  | 50 - 59 | 40 (4.04) | 11 (2.46) |  | 0.73 | 0.368 - 1.45 | 0.470 |
|  | 60 - 69 | 13 (1.31) | 7 (1.56) |  | 3.48 | 1.66 - 7.29 | 0.002 |
|  | >69 | 0 (0.00) | 1 (0.22) |  | 6.65 | 0.17 - 260.90 | 0.692 |
| Gender |  |  |  |  |  |  |  |
|  | Male | 555 (56.06) | 199 (44.52) | <0.001 | 1.00 | - | - |
|  | Female | 431 (43.53) | 246 (55.03) |  | 1.59 | 1.27 - 1.99 | <0.001 |
|  | *Prefer not to say | 4 (0.40) | 2 (0.45) |  | - | - | - |
| Education |  |  |  |  |  |  |  |
|  | No formal education | 0 (0.00) | 2 (0.44) | 0.045 | 1.00 | - | - |
|  | High School | 36 (3.63) | 24 (5.37) |  | 0.13 | 0.004 - 4.13 | 0.2504 |
|  | College (Bachelor) | 637 (64.34) | 260 (58.16) |  | 0.08 | 0.003 - 2.44 | 0.1186 |
|  | Masters | 219 (22.12) | 104 (23.26) |  | 0.09 | 0.003 - 2.85 | 0.1501 |
|  | Ph.D. | 56 (5.65) | 35 (7.82) |  | 0.13 | 0.004 - 3.82 | 0.2265 |
|  | Others | 42 (4.24) | 22 (4.92) |  | 0.10 | 0.003 - 3.25 | 0.1813 |
| Background |  |  |  |  |  |  |  |
|  | Non - Scientific/<br>Non - Medical | 367 (37.07) | 218 (48.76) | <0.001 | 1.00 | - | - |
|  | Scientific/Medical | 623 (62.92) | 229 (51.24) |  | 0.62 | 0.49 - 0.78 | <0.001 |
| Nationality |  |  |  |  |  |  |  |
|  | Egypt | 86 (8.68) | 219 (48.99) | <0.001 | 1.00 | - | - |
|  | Nigeria | 904 (91.32) | 228 (51.11) |  | 0.09 | 0.07, 0.13 | <0.001 |

$\chi^2$ - chi square; DF - degree of freedom; OR - odds ratio; CI: confidence interval; \* - excluded from the multivariable logistic regression analysis

The level of education, background, and nationality greatly affected the perception of global and community response to curbing the spread of COVID-19 and preventing the occurrence of any future pandemic. Educated respondents were 2.58 (95% CI: 0.09 -77.55; p > 0.999) to 6.54 (95% CI: 0.21 - 202.40; p = 0.543), more likely to have positive perceptions of the global responses than non-educated respondents. Similar to the attitude, scientific/medical respondents were 1.6× (95% CI: 0.56 - 0.87; p < 0.001) more likely to have better perceptions of the global responses than those with non-scientific/non-medical background (Table 6).

**Table 6.** Analysis of demographic characteristics as factors influencing the perceptions of respondents from Nigeria and Egypt towards COVID-19 pandemic.

| Independent variables | | Satisfactory (%) | Unsatisfactory (%) | p -value ($\chi^2$) | OR | 95%CI | p -value |
| --- | --- | --- | --- | --- | --- | --- | --- |
| Age |  |  |  |  |  |  |  |
|  | 18 - 29 | 439 (49.21) | 267 (48.99) | 0.888 | - | - | - |
|  | 30 - 39 | 301 (33.74) | 190 (34.86) |  | - | - | - |
|  | 40 - 49 | 104 (11.65) | 64 (11.74) |  | - | - | - |
|  | 50 - 59 | 35 (3.92) | 16 (2.93) |  | - | - | - |
|  | 60 - 69 | 12 (1.31) | 8 (1.47) |  | - | - | - |
|  | >69 | 1 (0.11) | 0 (0.00) |  | - | - | - |
| Gender |  |  |  |  |  |  |  |
|  | Male | 473 (53.02) | 281 (51.55) | 0.732 | - | - | - |
|  | Female | 416 (46.63) | 261 (47.88) |  | - | - | - |
|  | Prefer not to say | 3 (0.33) | 3 (0.55) |  | - | - | - |
| Education |  |  |  |  |  |  |  |
|  | No formal education | 2 (0.22) | 0 (0.00) | 0.03 | 1.00 | - | - |
|  | High School | 26 (2.91) | 34 (6.23) |  | 6.54 | 0.21 - 202.40 | 0.543 |
|  | College (Bachelor) | 553 (61.99) | 344 (63.11) |  | 3.11 | 0.10 - 92.94 | 0.95 |
|  | Masters | 213 (23.87) | 110 (20.18) |  | 2.58 | 0.09 - 77.55 | >0.9999 |
|  | Ph.D. | 57 (6.39) | 34 (6.23) |  | 2.98 | 0.09 - 91.26 | 0.984 |
|  | Others | 41(4.59) | 23 (4.22) |  | 2.81 | 0.09 - 86.84 | >0.999 |
| Background |  |  |  |  |  |  |  |
|  | Non - Scientific/<br>Non - Medical | 334 (37.44) | 251(46.05) | 0.001 | 1.00 | - | - |
|  | Scientific/Medical | 558 (62.56) | 294 (53.95) |  | 0.69 | 0.56 - 0.87 | 0.001 |
| Nationality |  |  |  |  |  |  |  |
|  | Egypt | 165 (18.49) | 140 (25.68) | 0.001 | 1.00 | - | - |
|  | Nigeria | 727 (81.51) | 405 (74.32) |  | 0.66 | 0.51 - 0.85 | 0.002 |

254 $\chi^2$ - chi square; DF - degree of freedom; OR - odds ratio; CI: confidence interval.

## Discussion

To the best of our knowledge, this research is one of the first studies examining the knowledge, attitude, and perceptions (KAP) toward COVID-19 in two of the most populated countries in Africa, Nigeria, and Egypt. Both countries announced the occurrence of their first COVID-19 cases in February, 2020 (12; 17; 18). Since then, the number of cases has increased with over 22,000 confirmed cases and over 900 deaths (10; 12).

Most of the respondents (62%) had a satisfactory knowledge level of the disease and the preventive measures against it. This is because both countries have a well - educated population (bachelor/master’s degree holders), mostly between 18 to 39 years (83%), and an average knowledge score of 74% indicated that most respondents were knowledgeable on COVID-19. It is also possible that the seriousness of the global pandemic in addition to daily updated from public health agencies in respective countries would have prompted the need to learn and acquire knowledge on COVID-19. However, this score is lower to the previous KAP studies on COVID-19 in China and Iran in which participants had an overall knowledge score of 90% (13; 19)

The internet (social media platforms- 84%) and TV (44%) were the main source of information for the participants. This is similar to the report by Abdelhafiz AS, et al. (20) where Facebook was the main source of information for young adults in their survey in Egypt. The internet (social media platforms) and TV had proved helpful for respondents to adapt with the physical social restraints during the COVID-19 compulsory lockdown in Nigeria and Egypt. In addition, almost half of our respondents (49%) were very satisfied with the social media coverage of the pandemic. This is lower than the 67% satisfaction rating of the social media coverage reported in Egypt (20). On the contrary, Roy et al. (21) reported 67% of Indians felt worried after receiving social media updates on the global burden of COVID-19.

The significant associations (p < 0.05) observed in this study between age, education, nationality, and background and the knowledge score of COVID-19 (Table 4) were similar to reports from other KAP studies from China, Egypt, and India in which participants who were well educated, young age or with high socioeconomic level had better knowledge of COVID-19 than the others (13; 20; 21).

Although this study was conducted during the compulsory lockdown in both countries, the optimistic attitude of Egyptians and Nigerian could be seen in a mean attitude score of 6.9 ± 1.2. Most (67%) of the respondents had generally satisfactory attitudes, recognizing the importance of social distancing (96%), and following the health recommendations (92.5%). However, only 36% followed all the recommendations. This might be due to the severe economic hardship faced by the citizens of both countries associated with workers who need to earn their daily wages, and poor government palliative plans for the citizens. This is further buttressed by the fact that only 39% of the respondents were convinced that their governments have done enough to curb the spread of the SARS-CoV-2. This distrust in the management of the pandemic might also be due to the low testing capability, and lack of strict enforcement of the compulsory lockdown. More so, in many African countries, reports of porous borders, congested cities, increased hunger and poverty, poor health literacy, and expensive face masks and hand sanitizers have all been obstacles against control measures (Lucero - Prisno DE, et al., 2020).

All of the respondents agreed on the importance of handwashing and other preventive measures in reducing the chances of being infected. A similar positive attitude towards most preventive measures were earlier reported in India (21) and Egypt (20) but the later noted some reluctancy in following some recommendations such as the use of a face mask. In another study conducted in China, most of the participants followed the health recommendations and less than 4% went to crowded places or went outside without a facemask. Chinese were also optimistic about the success of their COVID-19 control program (13). In our study, 96% of respondents considered self-isolation essential and effective, hence avoiding places with confirmed COVID-19 cases. This finding may support the lower number of recorded cases initially observed in Egypt and Nigeria. Comparably, in a KAP study conducted by Chan et al. (2015) on the H7N9 influenza pandemic, most respondents did not take the seasonal influenza as serious as 42.3% of the respondents did not avoid going to places that had the H7N9 confirmed cases.

While some participants were bored (52%), nervous/anxious (47%), afraid (44%) and stressed (30%). Others felt optimistic (18%) and happy (1.4%). Sixty - six (66%) of Nigerians and Egyptians were highly optimistic that collectively, the world can reduce the impact or prevent the occurrence of a similar future pandemic. This attitude is encouraging as it would facilitate eventual control of the pandemic.

Only 25.4% of the respondents were not satisfied with the WHO’s handling of the global pandemic. This high rating of the WHO’s efforts at coordinating global health might be attributed to the daily disease burden updates, press conferences, provision of authentic information, travel advise, and support for the health authorities of both countries.

It was remarkable that most of the participants acknowledged the importance of in-depth scientific research in areas of vaccines and diagnostics; and the need for increased multi - sectoral collaborations (on human, animal and environmental health) using the one health approach as measures that can help prevent the occurrence of a future pandemic.

The major limitations of this study were the low internet penetration rate in Nigeria (42%) and Egypt (54%); in which a significant proportion of the population could not gain access to this online survey. This, coupled with the lockdown limited the sample size of this preliminary study to 1437 (Nigeria - 1132 and Egypt - 305). A more encompassing global survey is currently being undertaken. Also, the data was skewed in favor of young respondents (18 – 39 years) due to their profound interest in social media. Our results cannot be generalized for Africa as a whole as each country had specific measures and peculiarities with regards to controlling the pandemic. For example, in Nigeria and Egypt, not all states have closed their borders, permitting the free movement of people across states.

## Conclusion

The COVID-19 pandemic has profound medical, economic, and psycho-social effects, with over 300,000 lives lost globally. Assessing the KAP of respondents and further education of the general public has proved effective in changing risk perception of the populace and resulted in attitudinal changes that were necessary to reduce the epidemic disease burden (23). Adequate monitoring of social media platforms to confirm and improve the quality of information delivered to the people is of prime importance (24).

Both Nigerians and Egyptians have a good knowledge of the pandemic and have a satisfactory attitude and perceptions towards the global response. However, we recommend increased adherence to the health regulations of both countries. Similarly, mental health support should be made more readily available to the populace. Both governments need to strengthen their health systems, and improve their surveillance activities, to be able to estimate and detect cases, trace contacts, properly isolate infected patients, and effectively apply standard infection prevention and control measures. In addition, they should continuously provide accurate and timely information to their masses.

## Data Availability

The Data is available on as supplementary data.

## Declarations

### • Availability of data and materials

The survey instrument and dataset are available as supplementary data.

## • Acknowledgments

We acknowledge Stephanie Germon for validating the survey instrument. We equally acknowledge the dedication of our friends in sharing the questionnaire on social media.

## • Authors’ contributions

EH, AZ and AIA planned the study. EH, OI and AIA contributed equally to the study. All co - authors participated in data collection. EH, OI, OO, and AIA drafted the manuscript. EH, OI, OB, AZ, OO, and AIA did the overall review of the manuscript. All authors read and approved the final study.

## Supplementary data

**Table s1:**
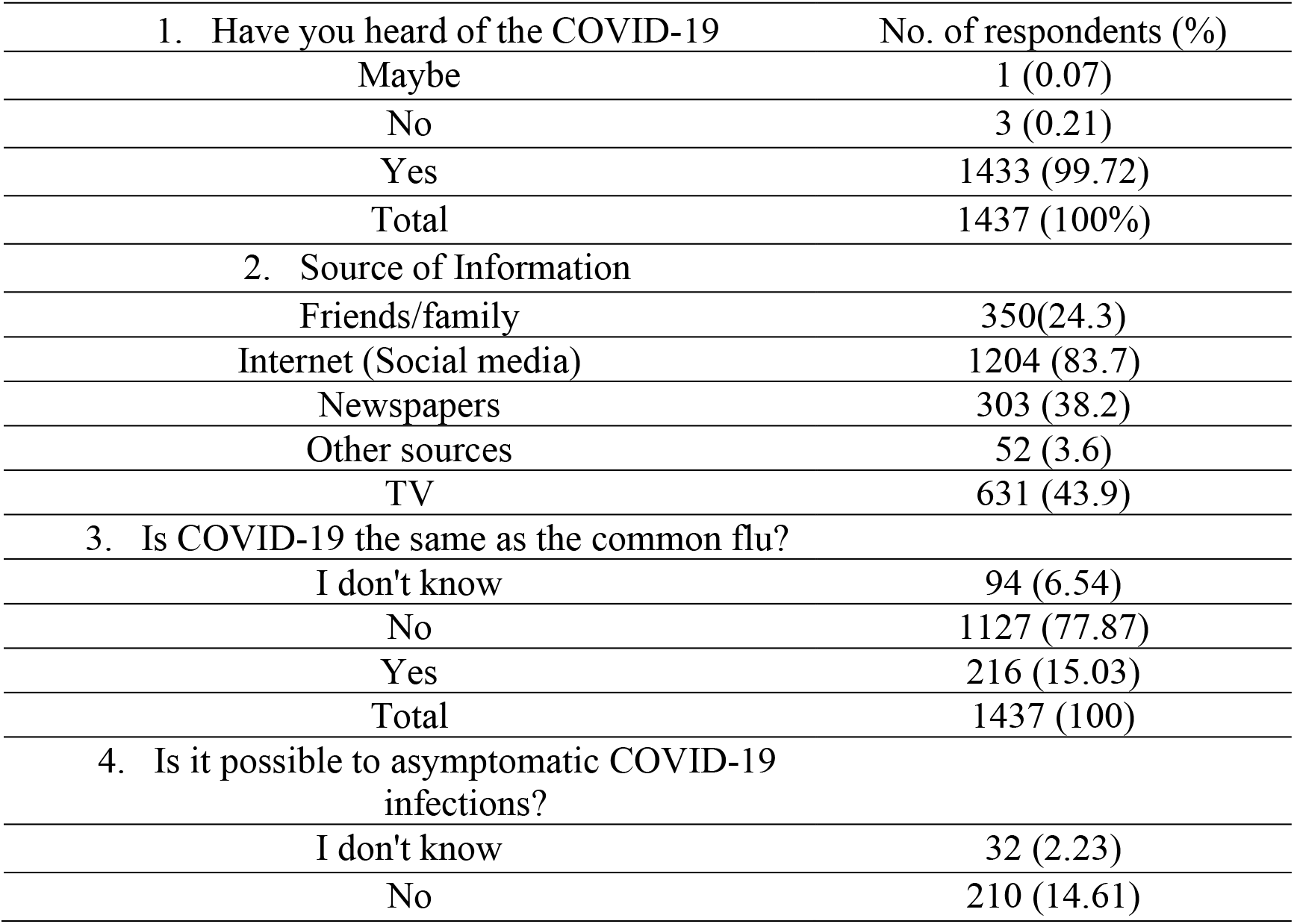
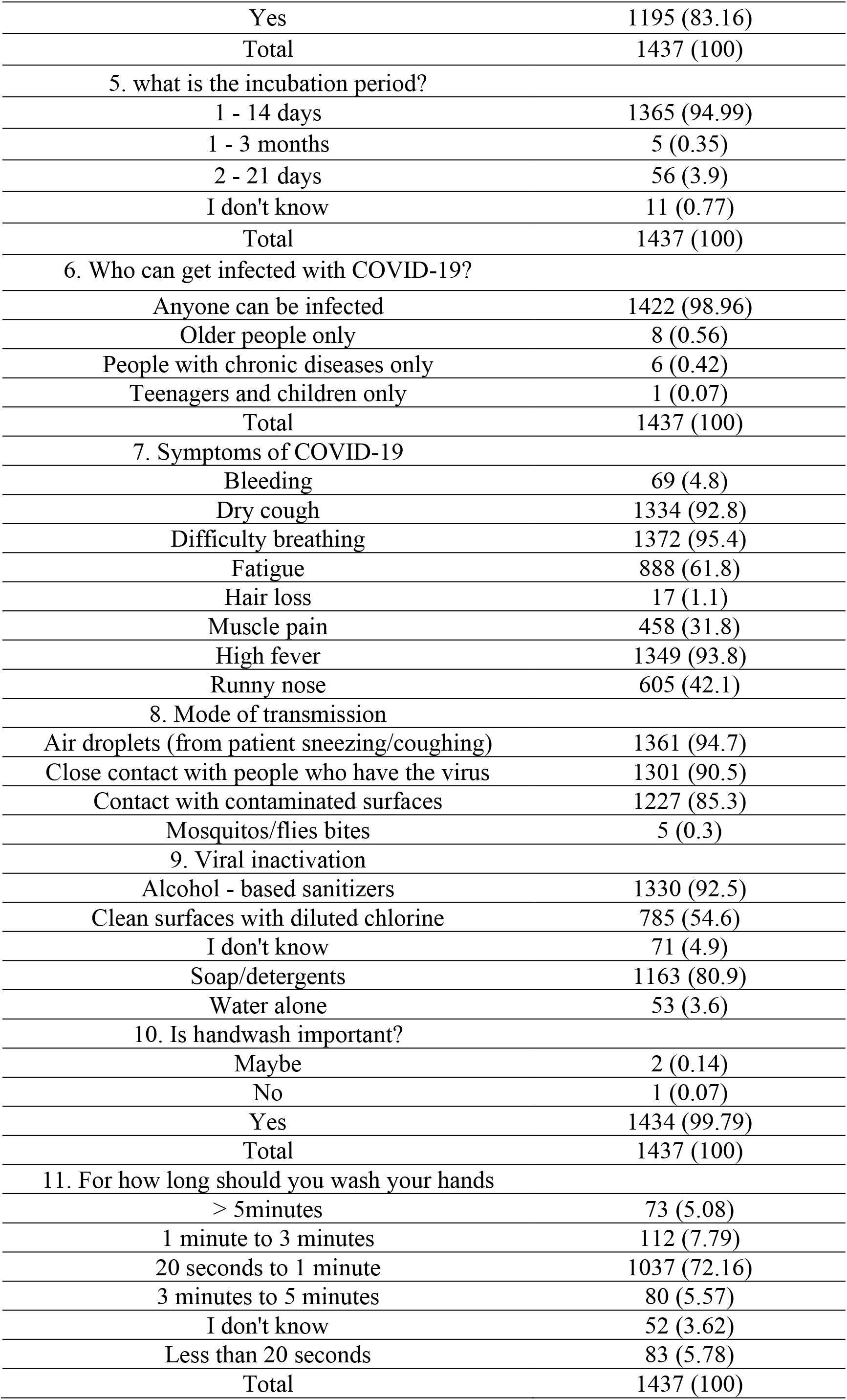
Descriptive statistics (Correct answer rate) of knowledge of COVID-19 pandemic in Nigeria and Egypt.

**Table s2.**
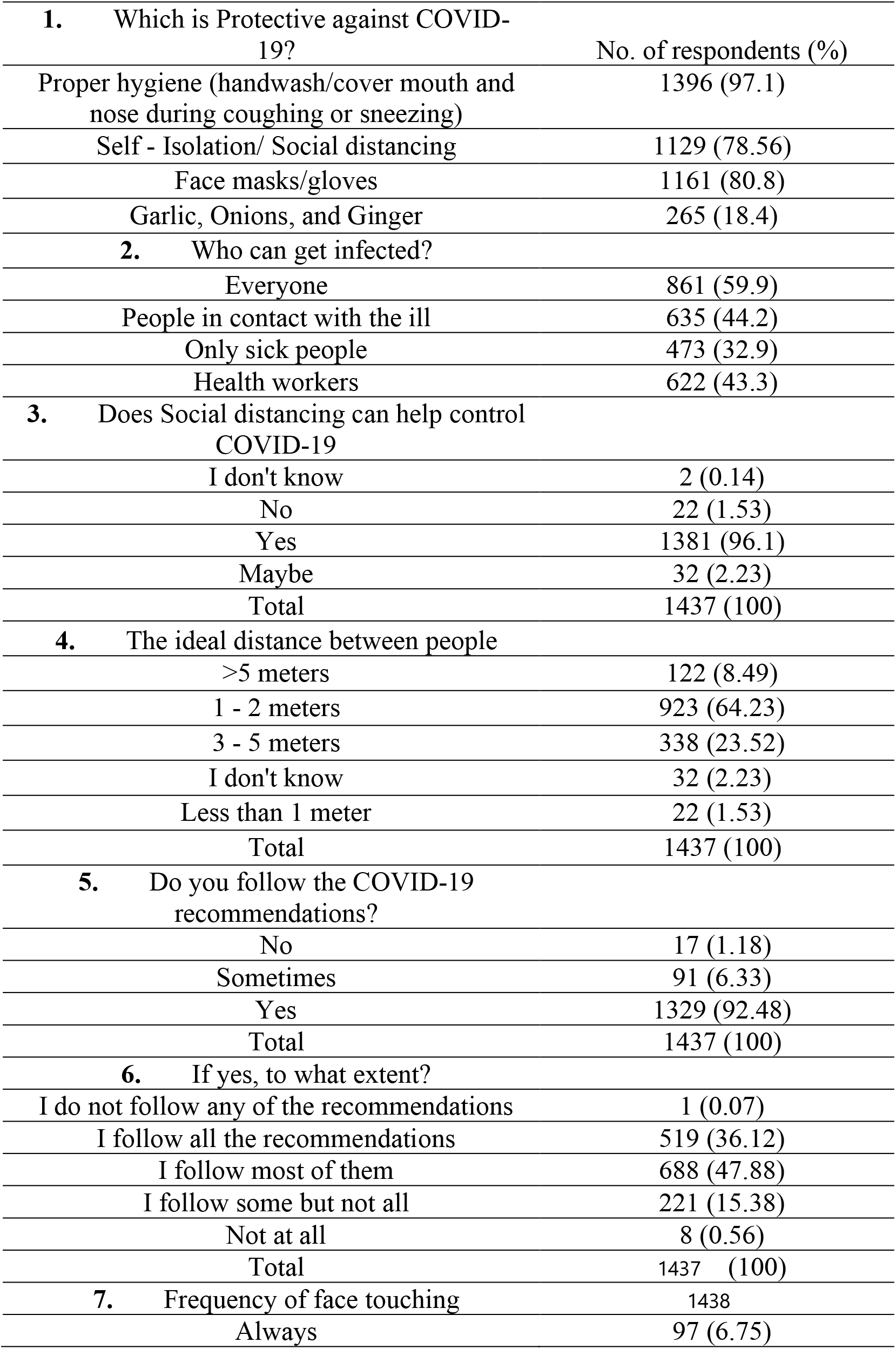
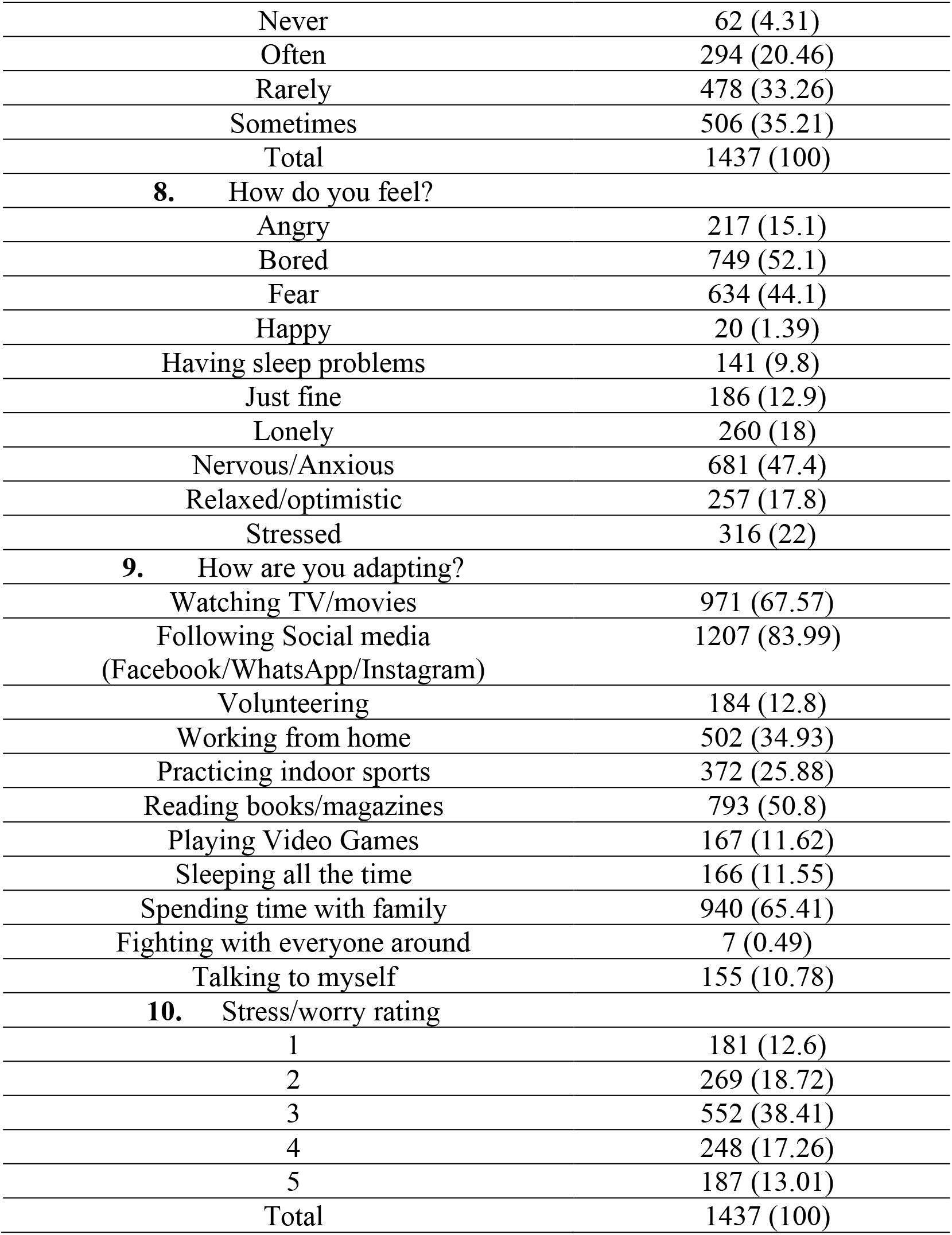
Descriptive statistics (Correct answer rate) of attitude towards preventive measures to the COVID-19 pandemic in Nigeria and Egypt.

| 1. Which is Protective against COVID-19? | No. of respondents (%) |
| --- | --- |
| Proper hygiene (handwash/cover mouth and nose during coughing or sneezing) | 1396 (97.1) |
| Self - Isolation/ Social distancing | 1129 (78.56) |
| Face masks/gloves | 1161 (80.8) |
| Garlic, Onions, and Ginger | 265 (18.4) |
| 2. Who can get infected? |  |
| Everyone | 861 (59.9) |
| People in contact with the ill | 635 (44.2) |
| Only sick people | 473 (32.9) |
| Health workers | 622 (43.3) |
| 3. Does Social distancing can help control COVID-19 |  |
| I don't know | 2 (0.14) |
| No | 22 (1.53) |
| Yes | 1381 (96.1) |
| Maybe | 32 (2.23) |
| Total | 1437 (100) |
| 4. The ideal distance between people |  |
| >5 meters | 122 (8.49) |
| 1 - 2 meters | 923 (64.23) |
| 3 - 5 meters | 338 (23.52) |
| I don't know | 32 (2.23) |
| Less than 1 meter | 22 (1.53) |
| Total | 1437 (100) |
| 5. Do you follow the COVID-19 recommendations? |  |
| No | 17 (1.18) |
| Sometimes | 91 (6.33) |
| Yes | 1329 (92.48) |
| Total | 1437 (100) |
| 6. If yes, to what extent? |  |
| I do not follow any of the recommendations | 1 (0.07) |
| I follow all the recommendations | 519 (36.12) |
| I follow most of them | 688 (47.88) |
| I follow some but not all | 221 (15.38) |
| Not at all | 8 (0.56) |
| Total | 1437 (100) |
| 7. Frequency of face touching | 1438 |
| Always | 97 (6.75) |

|  |  |  |
| --- | --- | --- |
|  | Never | 62 (4.31) |
|  | Often | 294 (20.46) |
|  | Rarely | 478 (33.26) |
|  | Sometimes | 506 (35.21) |
|  | Total | 1437 (100) |
| **8.** | How do you feel? |  |
|  | Angry | 217 (15.1) |
|  | Bored | 749 (52.1) |
|  | Fear | 634 (44.1) |
|  | Happy | 20 (1.39) |
|  | Having sleep problems | 141 (9.8) |
|  | Just fine | 186 (12.9) |
|  | Lonely | 260 (18) |
|  | Nervous/Anxious | 681 (47.4) |
|  | Relaxed/optimistic | 257 (17.8) |
|  | Stressed | 316 (22) |
| **9.** | How are you adapting? |  |
|  | Watching TV/movies | 971 (67.57) |
|  | Following Social media<br>(Facebook/WhatsApp/Instagram) | 1207 (83.99) |
|  | Volunteering | 184 (12.8) |
|  | Working from home | 502 (34.93) |
|  | Practicing indoor sports | 372 (25.88) |
|  | Reading books/magazines | 793 (50.8) |
|  | Playing Video Games | 167 (11.62) |
|  | Sleeping all the time | 166 (11.55) |
|  | Spending time with family | 940 (65.41) |
|  | Fighting with everyone around | 7 (0.49) |
|  | Talking to myself | 155 (10.78) |
| **10.** | Stress/worry rating |  |
|  | 1 | 181 (12.6) |
|  | 2 | 269 (18.72) |
|  | 3 | 552 (38.41) |
|  | 4 | 248 (17.26) |
|  | 5 | 187 (13.01) |
|  | Total | 1437 (100) |

**Table s3:**
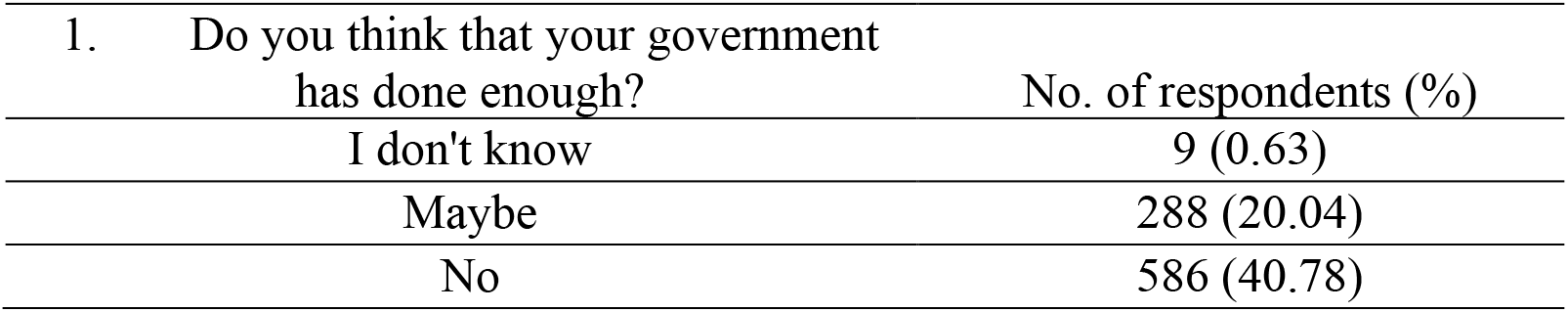
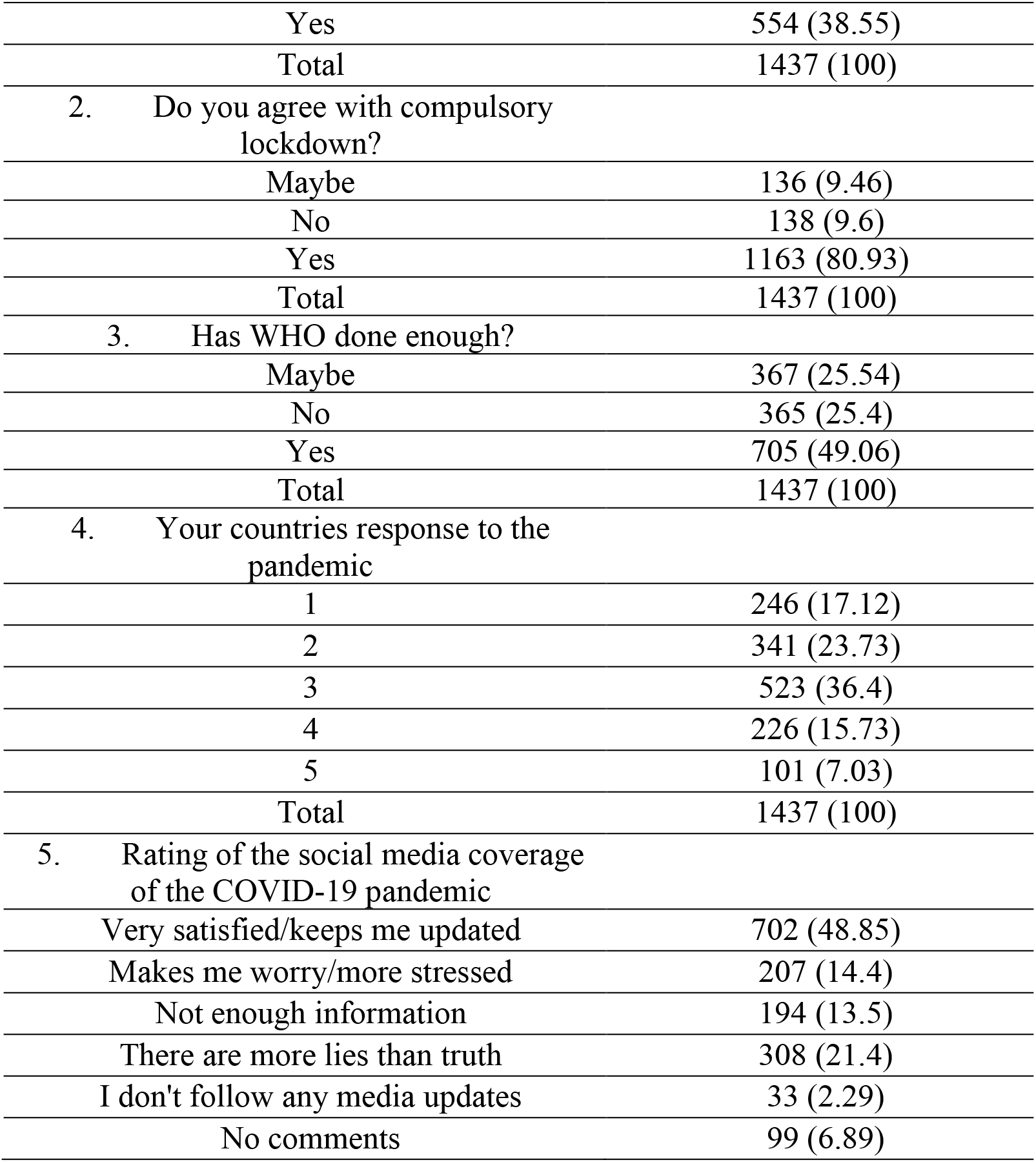
Descriptive statistics (Correct answer rate) of perception of the global response to the COVID-19 pandemic in Nigeria and Egypt.

**Table s4:** Descriptive statistics (Correct answer rate) of respondents to community response associated with the prevention of a future pandemic.I don’t know

|  |  |  |
| --- | --- | --- |
| 1. | What can a community do to reduce the spread? | No. of respondents (%) |
|  | Follow/respect the Health recommendations of my country | 1340 (93.24) |
|  | Eat Healthy/ Practice sports | 823 (57.27) |
|  | Social distancing/Avoid the crowd | 1277 (88.86) |
|  | Volunteer to support whenever possible | 731 (50.86) |
|  | Avoid handshakes and face kissing | 1170 (81.41) |
|  | Attending religious gatherings | 21 (1.46) |
|  | I don't know | 7 (0.49) |
| 2. | Can we prevent future pandemics? |  |
|  | I don't know | 22 (1.53) |
|  | Maybe | 259 (18.02) |
|  | No | 211 (14.68) |
|  | Yes | 945 (65.76) |
|  | Total | 1437 (100) |
| 3. | Preventing future global pandemic |  |
|  | Reduce international travels | 472 (32.8) |
|  | Establish early alerts and global warning systems for infectious diseases | 1175 (81.8) |
|  | Collaboration between environmental, animal and human health workers | 912 (63.5) |
|  | Intensify research on preventive measures such as vaccines/diagnosis | 1156 (80.4) |
|  | Improve surveillance in the human and animal health sectors | 1044 (72.65) |
|  | Raise public awareness of proper hygiene/healthy habits | 1030 (71.6) |
|  | Prioritize human life/health welfare over the animal or environmental ones | 340 (23.6) |
| 4. | Willingness to share info |  |
|  | Maybe | 86 (5.98) |
|  | No | 49 (3.41) |
|  | Yes | 1302 (90.61) |
|  | Total | 1437 (100) |

